# Scrutinising the COVID-19 data on 590.000 cases. A retrospective, population-based descriptive study for data quality surveillance and a review at 4.540.000 cases

**DOI:** 10.1101/2020.05.26.20113316

**Authors:** Oriol Gallemí Rovira

## Abstract

**Background:** Reports on the detected positive patients with COVID-19 are as per today the best estimation of a country spread of the pandemic. In order to evaluate the early indicators for true lethality and recovery time, the data where the model is built must be quality checked. Each country sets different procedures and criteria for fatality count due to COVID-19 and the health system is stressed by having insufficient testing, untracked patients and premature discharge. In this paper the dynamics behind such data quality issues are discussed throughout the disease course to support better modeling and decision-making processes in a stressed healthcare system.

**Methods:** Based on data compiled and relayed by the Johns Hopkins University, tracking COVID-19 over 590.000 patients (march 27^th^, 2020), the data is clustered and compared with discrete regression. Regression parameters are restricted by a time interval of 1 day and must be meaningful for the diagnostic (i.e. a fatality cannot occur before the patient displays symptoms). Cumulative infection curves are taken and built. Infection baseline is based on the country official declaration. Infection synthetic curves are built from the Fatality count and the Recovered patient count. The adjusted parameters are τ=time to fatality (days), δ=time to discharge of recovered patients (days) and φ=case fatality rate (CFR in per unit, P.U.). Therefore, the discharge rate (recovery rate) is forced to be (1-φ).

Using forward or backward formulas have no other influence than the time reference. In both circumstances, time from Onset and Symptoms are neglected and shall be added if such dates are to be plot. There is a gap of two weeks since exposure to Hospital Admission to detection and the earlier the diagnose is done, the better the outcome.

Cumulative figures are used to smoothen the deviation and to provide the best estimator possible at the present time. The delay factor allows to compare figures belonging to the same date of detection.

Fast, daily models which can be used and integrated to a filtering stage on the parameter estimator in a complex approach are left out of scope. Continuous models can also be used and interpolation among the data points is another source of noise to be considered, especially when counting methods are suddenly changing as it is the case with COVID-19.

Countries were grouped as found representative for methodology illustration purposes. Results are discussed and compared across the different groups and potential indicators of this behavior are drawn for further study.

**Findings:** From 593.291 cases in the sample, and its 7 representative groups, the recovery time and the local CFR are negatively correlated, having the highest fatality rates (21%, Spain) the countries with shorter recovery time (11 days, Spain). Also, CFR can be an indicator of Infection inconsistencies (i.e. South Korea, CFR 1%, Time to recovery 25 days).

At the review part, focus is made on the inconsistencies detected in Germany and South Korea datasets as well as the potential misfits on China and Spain.

Overall, the Time to Fatality ranges between 4 and 8 days, and the mean is of 6 days (South Korea, 7 days; Japan, 6days). Only Germany and France are detecting earlier than other countries and admit 10 days before fatality occurs.

To date, shortening hospital discharge times seem to lead to patient reinfections (COVID-19 positive), and studies are working on this line.

**Interpretation:** One simple explanation for the local CFR and Recovery time correlation is to define such rate as a measure of the healthcare system overload. Anomalous CFR indexes point to a stressed healthcare system. The higher the overload, the more focus on critical cases and hence the higher local CFR.

The COVID-19 intrinsic CFR is unlikely to change by a factor of 10x from countries with similar lifestyle, GDP per capita and health services (i.e. the Mediterranean Basin, Northern Europe, etc.). Because of this fact, early CFR measured before Healthcare system overwhelming (COVID-19 free flow) are considered to be more accurate than the measured CFR while the outbreak is still ongoing,

Finally, the synthetic Infection indexes may be a helpful indirect measure of the real population infection rate and also used for data quality audit. Any model built upon inconsistent data will be complex to explain and justify.

**Funding:** No specific funding is raised.

## Introduction

Beginning in December 2019, a cluster of cases of pneumonia with unknown cause was reported in Wuhan, in the Hubei province of China and by December 31^st^, the Chinese government raised its concerns to the WHO and closed the potential source, a trade market from Wuhan. On January 23^rd^, China declares a local lockdown and by January 25^th^ an extended lockdown with more restrictive measures in place. By January 30^th^, WHO does not consider to be a Public Health Emergency of International Concern(1). A novel virus form denominated SARS-CoV-2 is sequenced and found to be fast adapting to new species infection, being humans among its hosts which develop the denominated COVID-19 disease. WHO declares the pandemic status by March 11^th^ 2020(2). Since the Chinese alert, the number of cases has exhibited a pandemic profile worldwide with an estimated CFR above 2%, and a strong human-to-human transmission, weaker to human-mammal pets.

Research in context

### Evidence before this study

Before this study, we searched Google Scholar, Elsevier and Springer repositories until March 25^th^, 2020 for articles describing the COVID-19 clinical course, symptomatic features, prognosis and epidemic modeling. SARS and MERS keywords were also used to extend the search on useful articles and lessons learnt from the past outbreaks. Diverse data sources were found and the Johns Hopkins University repository on Github which was selected for its continuous efforts to refine and curate the data released.

### Added value of this study

We developed a tool to validate raw data quality. As collateral outputs, we have estimates of COVID-19 features as Time to Fatality, Time to Recovery and Case Fatality Rate as well as a minimum Infections estimator. Such indicators can be used to assess infected detection procedures, having a large population of over 590.000 detected infections worldwide.

Our findings emphasize the relevance of proper data collection in early stages of an outbreak and provide insights on the procedures during the expansion, to validate the healthcare measures in place and its effects, suggesting potential improvement paths and proposing further lines of study to support fast data-driven, effective and efficient decision-making under pressure.

### Implications of all the available evidence

COVID-19 has a fast cycle on elder and sensitive subjects, leading to sudden ARDS (Acute Respiratory Distress Syndrome) and fast death since. By including data validation in early stage, the healthcare system capacity can be quickly prepared for the disease, triggering the responses earlier.

We focused our expert research on data and modelling, in order to define a clinical course^(3)(4)(5)^ to feed an explainable and actionable numerical model and contrast different data sources and to assess both the data quality and the clinical course estimators.

Estimating the real number of infections is found to be of paramount relevance in order to stop COVID expansion and other estimators(6) under study can complement the minimum found with the explained method.

Our main goal is to support decision-making and to deliver open tools for procedure setup and early actuation.

## Methods

### Study design

Based on data compiled and relayed by the Johns Hopkins University, tracking COVID-19 over 590.000 cases (march 27^th^, 2020), the data is clustered and compared with discrete regression. For reference, the same method is also applied to selected cases on 4.54 M cases (may 14^th^, 2020). Regression parameters are restricted by a time interval of 1 day and must be meaningful for the diagnostic (i.e. a fatality cannot occur before the patient displays symptoms). Cumulative infection curves are taken and built. Infection baseline is based on the country official declaration. Infection synthetic curves are built from the Fatality count and the Recovered patient count. The adjusted parameters are τ=time to fatality (days), δ=time to discharge of recovered patients (days) and φ=case fatality rate (CFR in per unit, P.U.). Therefore, the discharge rate is forced to be (1-φ).

Estimating the case fatality rate (CFR) during an outbreak is a complex work as data is incomplete, inconsistent, delayed and biased. Once the outbreak is complete, the CFR best estimator is:

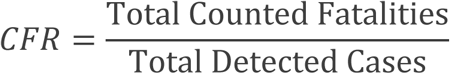

As the epidemic is still ongoing, estimators can be inconsistent and misleading as the data is strongly deformed by detection bias and delays:

1. Incomplete: Sample size is small to be not representative of a larger population. Studies of a few individuals deliver wide Confidence Intervals (CI) which make them insufficiently representative. To complete the sample, testing must be extended.
2. Inconsistent: Each hospital, province and state set different standards for prognosis and considers admissions and discharges upon a wide spectrum of diagnose.
3. Delayed: Admissions are accepted once symptoms become evident. Therefore, the incubation period is completed and beyond. Recovered patients are discharged upon symptomatic relief after hospitalization time, albeit the viral load may still be present in the recovered patient. Most fatalities occur in a different timeline from recovered.
4. Biased: Patients attending the hospital are mobility constrained. Elder and younger patients use to be accompanied, exposing young adults to infection. This is a potential source of biased sample demographics attended at the hospital with an over-representation of mid-aged patients over a true population pyramid. Another bias is that there is no control group in the general population and there is no way to precisely quantify the real spread of COVID-19 at the moment of writing.

To provide a meaningful CFR and Hospitalization Rate (HR), time and bias must be included in the time-count model. The reconstruction formulas for infections become (Eq.1) and (Eq.2):

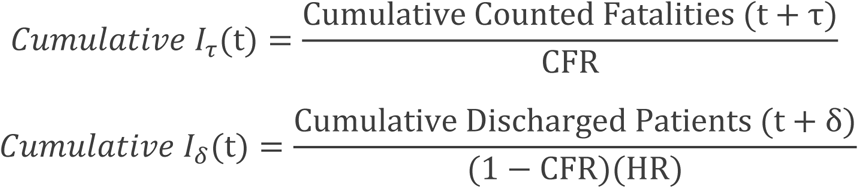

Using forward or retrospective formulas has no other influence than the time reference. In both circumstances, time from Onset and Symptoms are neglected and shall be added if the Onset date aims to be plot.

Cumulative figures are used to reduce the deviation and to provide the best estimator possible at the present time.

Delays allow to compare figures belonging to the same date of detection, regardless of their origin.

Fast, daily models which can be used and integrated to a filtering stage on the parameter estimator in a complex approach are left, out of scope. Continuous models can also be used and interpolation among the data points is another source of noise to be considered, especially when counting methods are suddenly changing as it is the case.

**Countries were grouped as found representative for methodology illustration purposes**. Results are discussed and compared across the different groups and potential indicators of this behavior are drawn for further study.

As the country with more tests conducted per capita is statistically closer to have a CFR in the order of magnitude of the IFR, an estimated minimum number of infections for the country *i* is computed by the use of the equation.

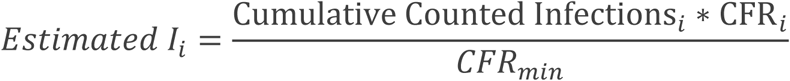

### Data Acquisition and processing

Consolidated data is taken from the JHU repository with the Countries’ cumulated Infected, Recovered and Fatality cases. From such and using the correction formulas and adjusted to an integer number of days, the values of τ=time to fatality (days), δ=time to discharge of recovered patients (recovery, days) and φ=case fatality rate are computed.

### Statistical Analysis

Preliminary filtering was done with Ms Excel 2016. Heuristic adjustments were done to reach R squared fit above 0,99 threshold. Use of discrete number of days (integer) to minimize noise was chosen over moving average windows or spline generation for interpolation.

Kaplan-Meier estimators are not used as the unknown infections is likely much higher and the method does nor appropriately work with censored values above 40%(7). To note, in Hong-Kong SARS, the estimated censored rate was of 86%. This implies a minimum factor of 6 to the minimum real infections figure.

### Role of the funding source

There was no funding source for this study. The author had access to the Johns Hopkins University repository(8) on Github and had the final responsibility for submission of the article.

## Results

593.291 patients positively diagnosed with COVID-19 are taken as a baseline population. The Infected curve is reconstructed from the Fatalities curve and from the Recovered (discharged) curve.

Country figures display a wide range of parameter magnitudes while the fit has a good adjustment. US has a Time to Fatality of 5 days and a Time to recover of 20 days with a CFR of 4%. Contagion is still growing and more datapoints are needed to properly reconstruct the curve from recovered patients.

**Figure 1.**
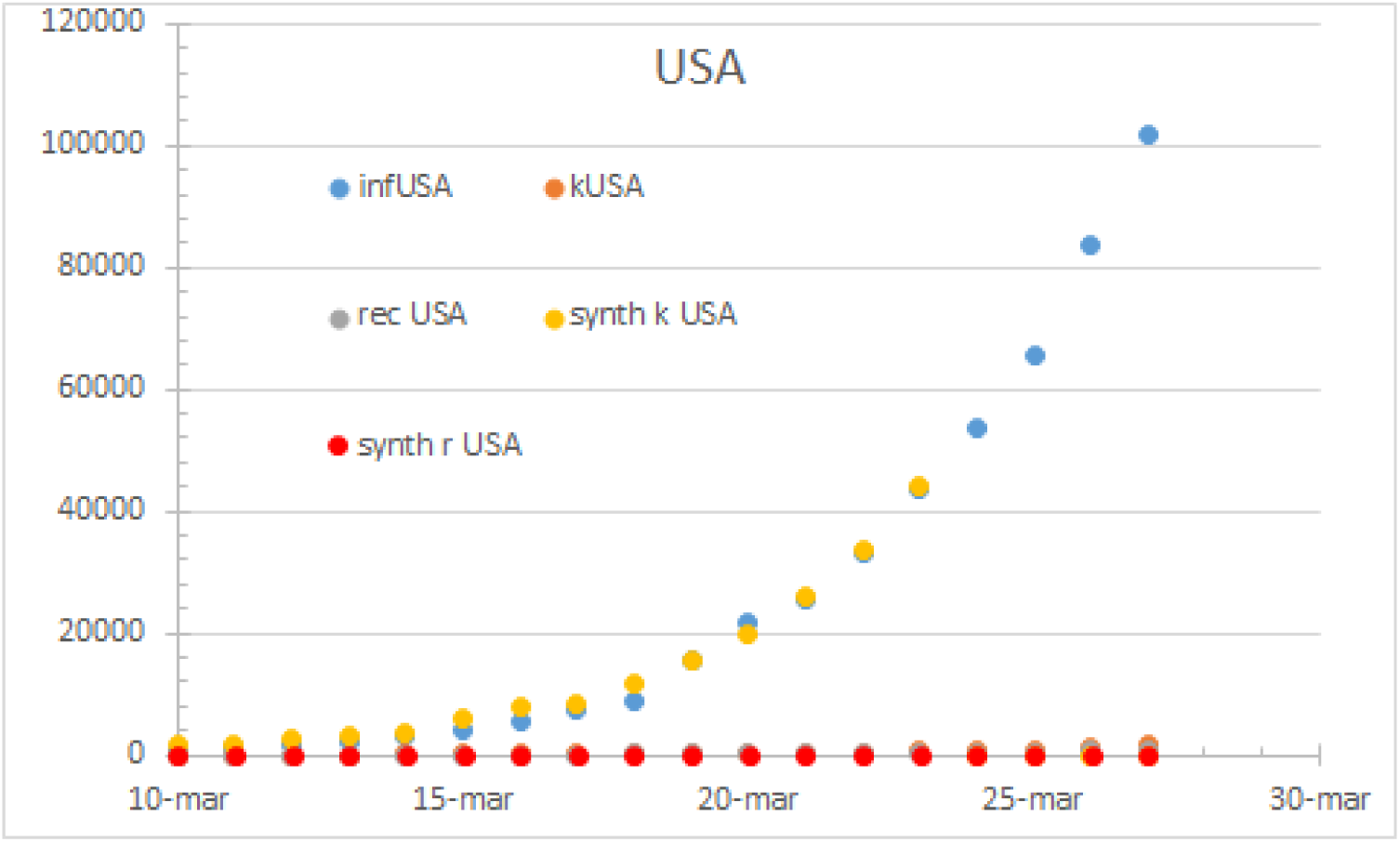
USA status as per may 24th. Note the missing recovered figures

**Figure 2.**
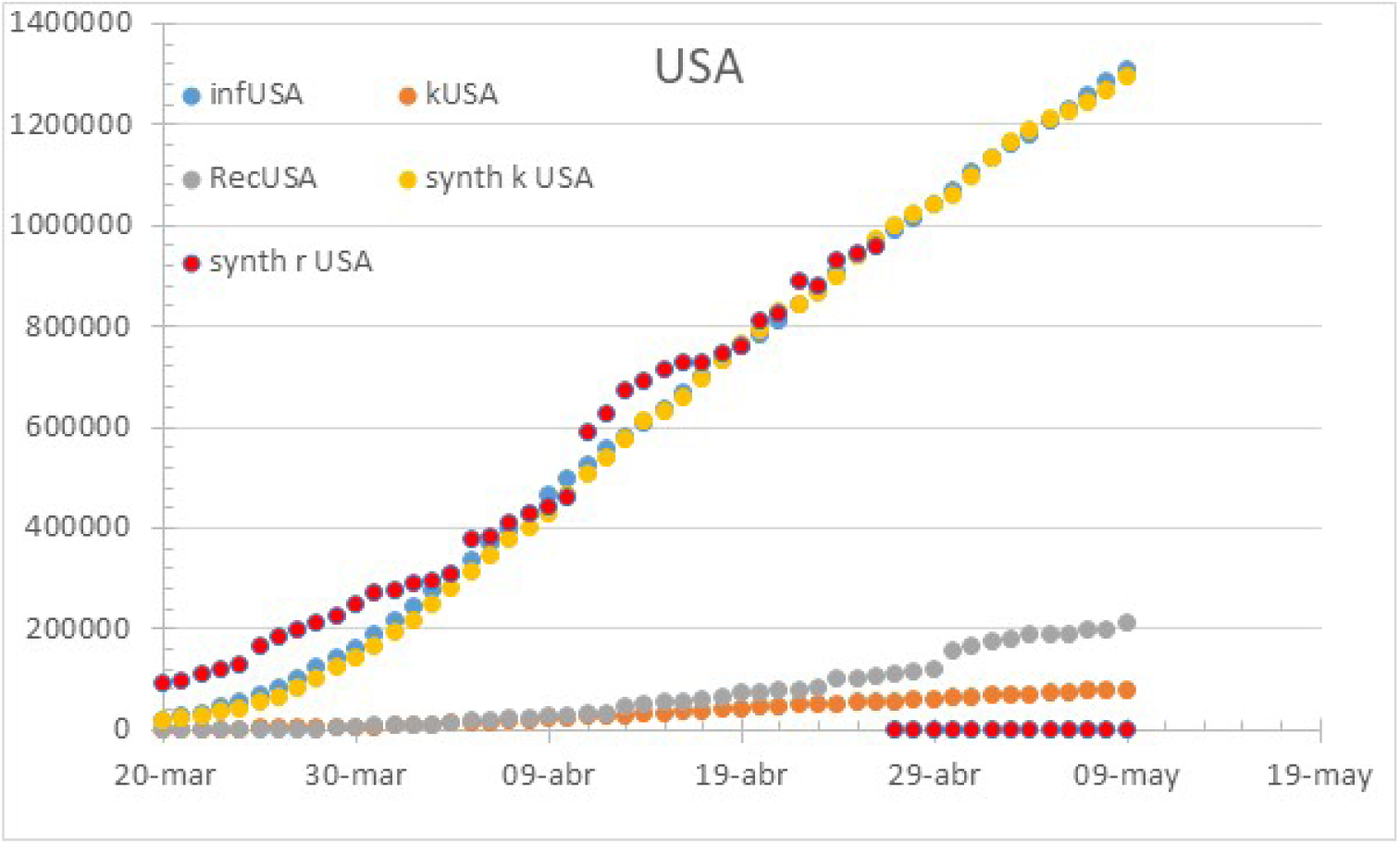
Status of the USA on may 14th. Note the 28% hospitalization rate on April 26th

China has a Time to fatality of 8 days and a Time to recover of 21 days with a CFR of 4%. Having a track record limited since the lockdown, the reconstructed curves for infected from recovered and fatalities match, and the synthetic ones have the best fit compared to the declared infections with a smoother evolution.

**Figure 3:**
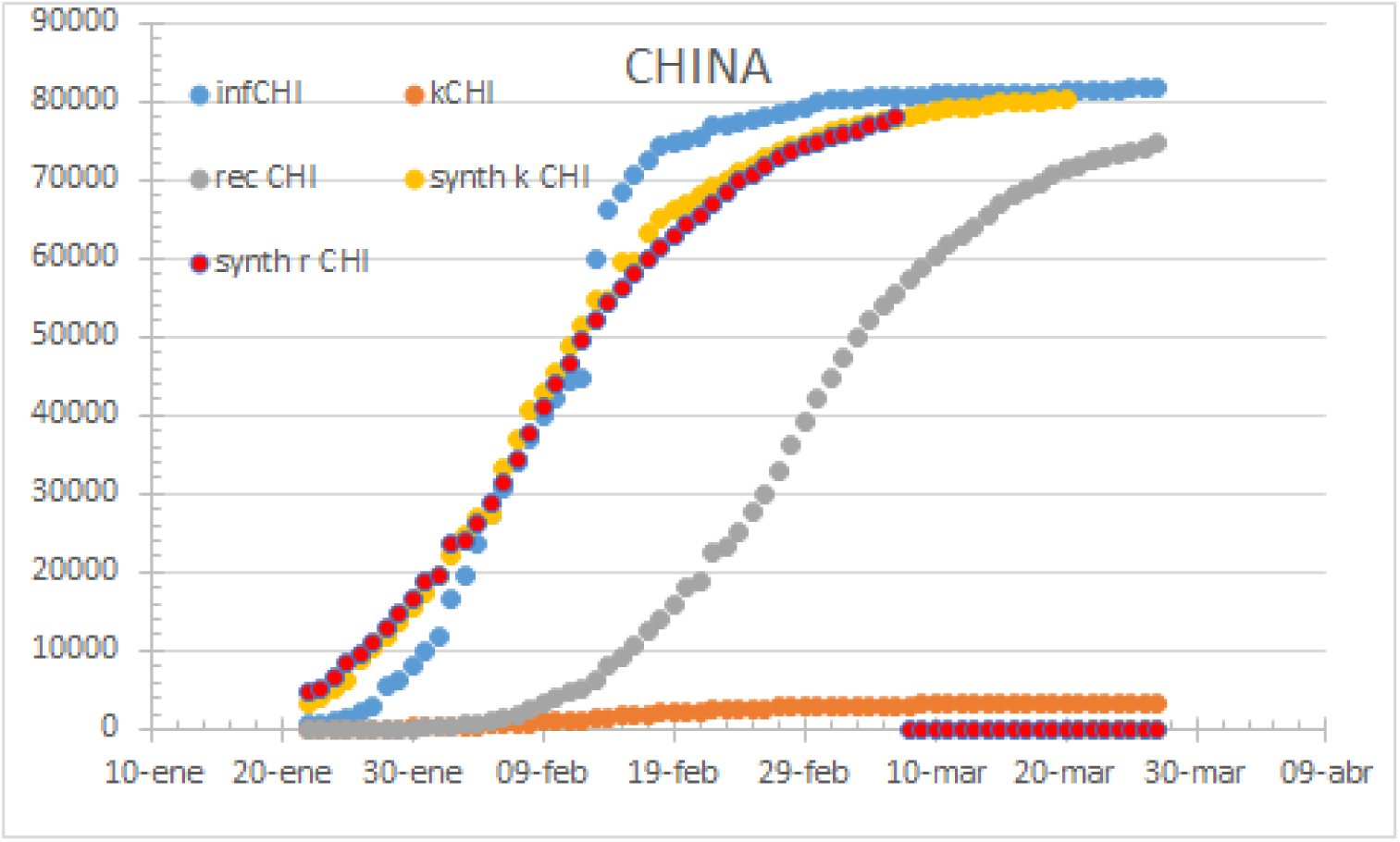
China cases followed a perfect 100% hospitalization rate flawless curve. Presented march 24th..

Europe’s Italy, Germany and France have a Time to fatality of 4 days and a Time to recover of 15 days with a CFR of 9%. Despite having a different policy, the aggregated Infection curve is matching the reconstructed one from both the fatality (yellow) and recovered (red).

**Figure 4.**
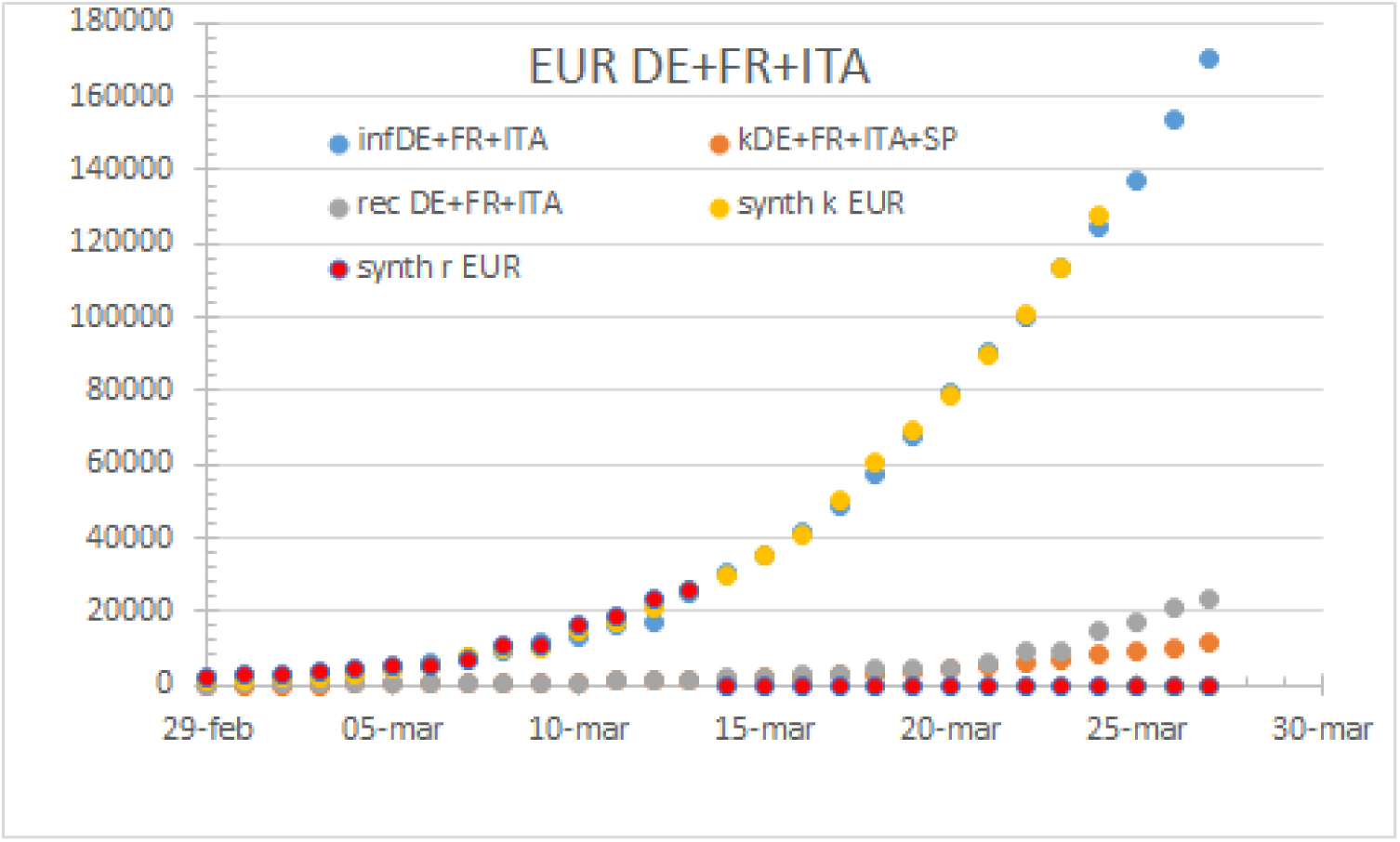
Europe by march 24th. Note the beginning of country bias.100% hospitalization rate.

The EU case requires a further analysis as countries started to modify its accounting pattern since march 24^th^. Therefore, Belgium is presented as a case with a 32% Hospitalization Rate, 7 days to fatality and 6 days to recovery.

**Figure 5.**
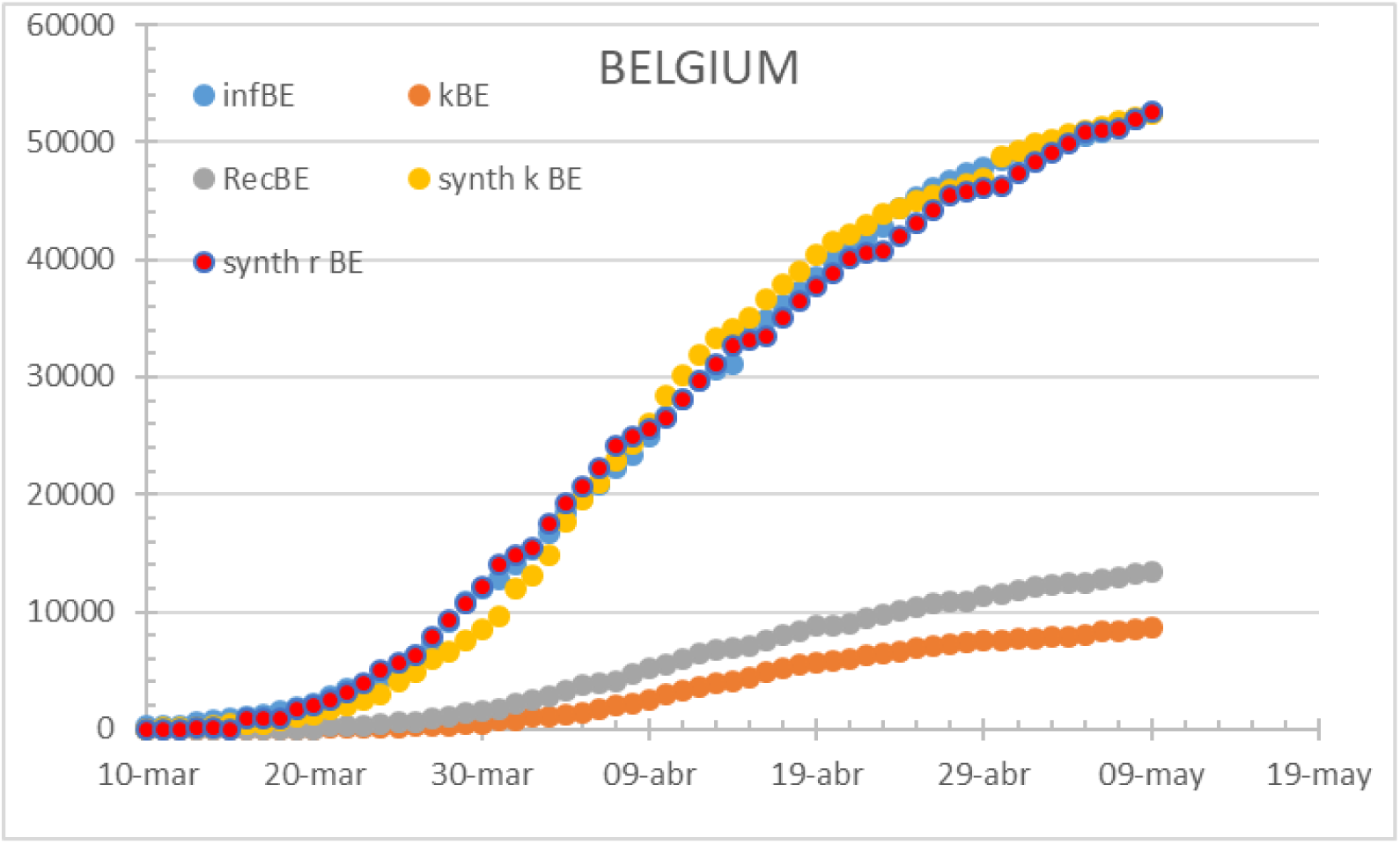
The Belgium case exhibits a fast path to diagnose which patients are more severely affected. May 14th

Italy has a perfect match on diagnose and fatalities and its Healthcare system became overwhelmed by mid-April reaching its absolute minimum HR. The model shows 5 days to fatality and 10 days of hospitalization to recovery.

**Figure 6.**
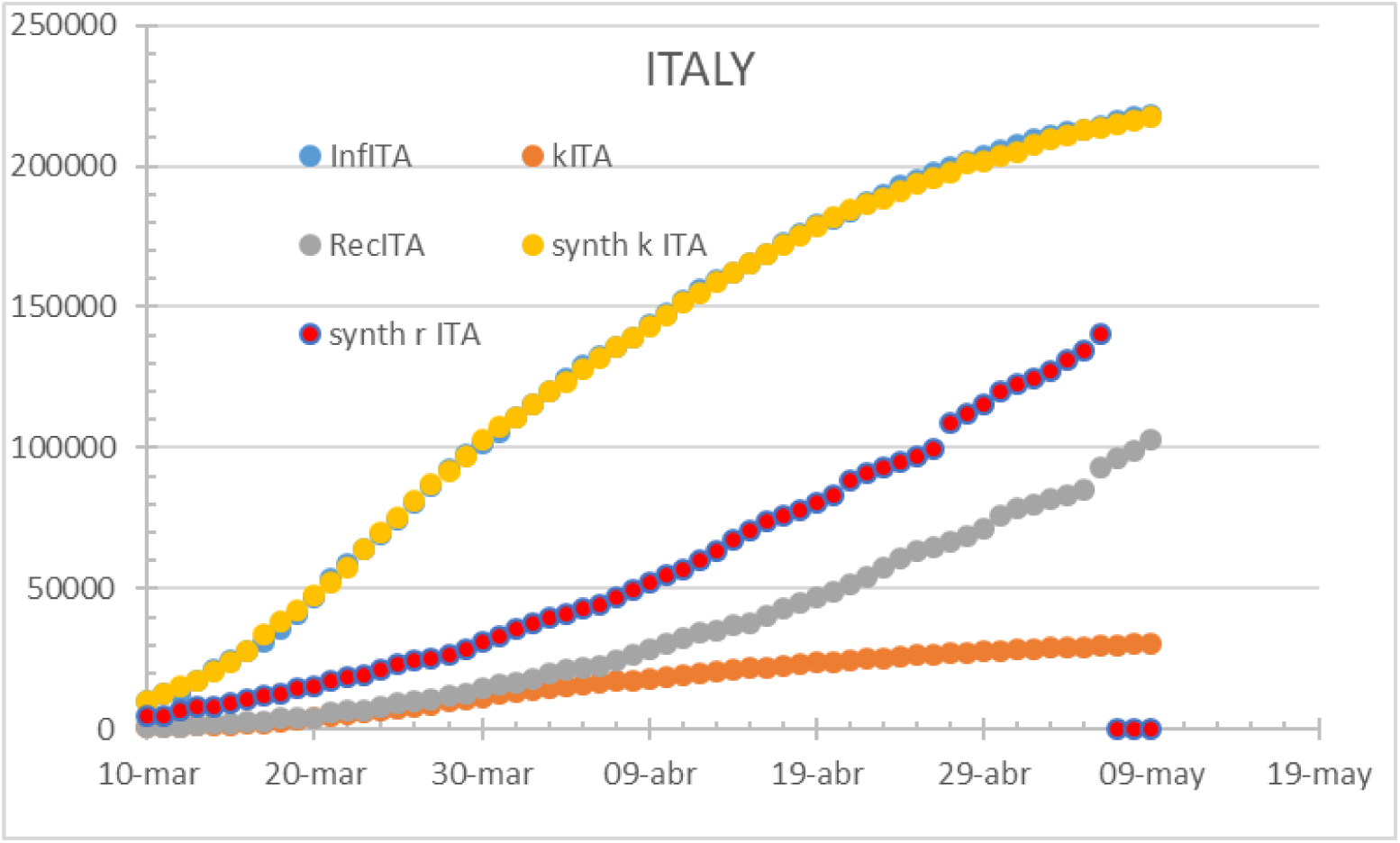
Italy on may 24th. To note the variation of HR along the infection, still below 70%

France has early detected gaps and pitfalls on its methodology and proceeded to correct and fix testing and accounting. The 7-day cycle is noticeable but still credible with an hospitalization rate of 41%, 10 days to fatality and 11 days to recovery as per may 14^th^.

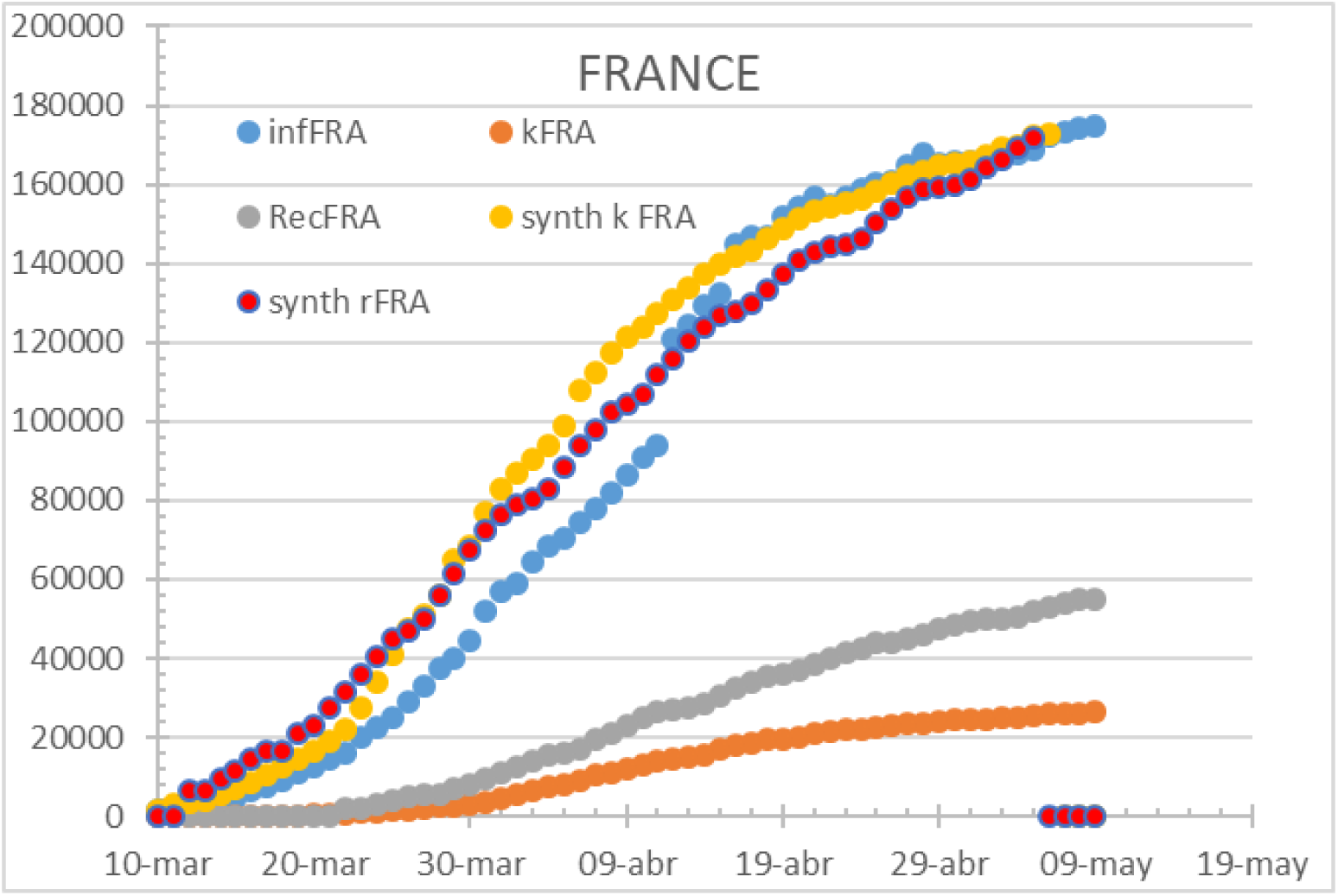

Spain has a Time to fatality of 7 days and a Time to recover of 11 days with a CFR of 21% by march 24^th^. Being all three curves consistent, it replicates the European fit, with the only differences of parametrization.

**Figure 7.**
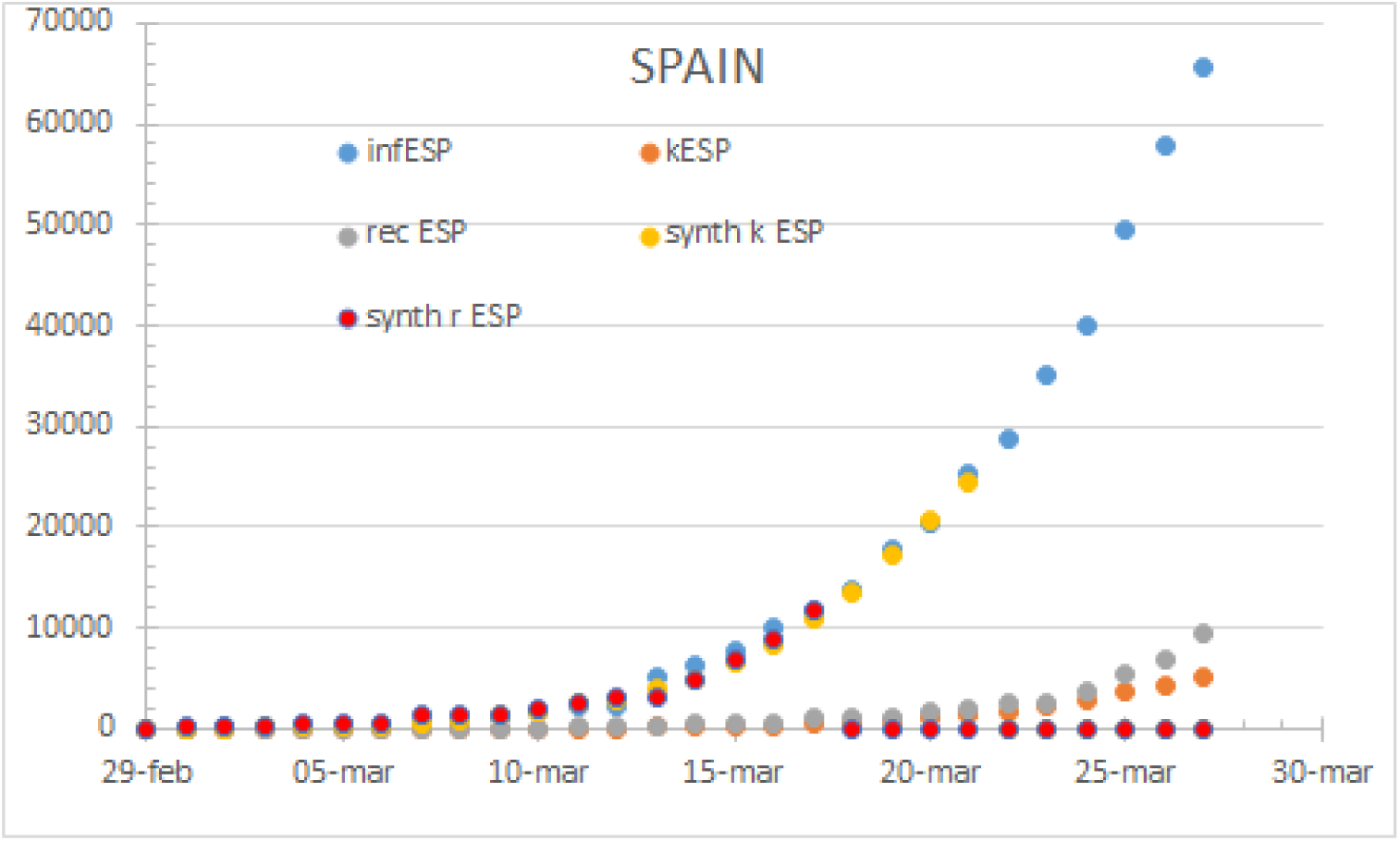
Spain by march 24th. Unbiased information delivers overlapping curves with a 100% hospitalization rate.

Spain has modified the testing, diagnose and accounting methods in several stages, which instead of matching the fatality curve as France, is forcing to match the infections curve since late April. The below exposed parameters include 67% HR, 3 days to fatality and 11 days to recovery.

**Figure 8.**
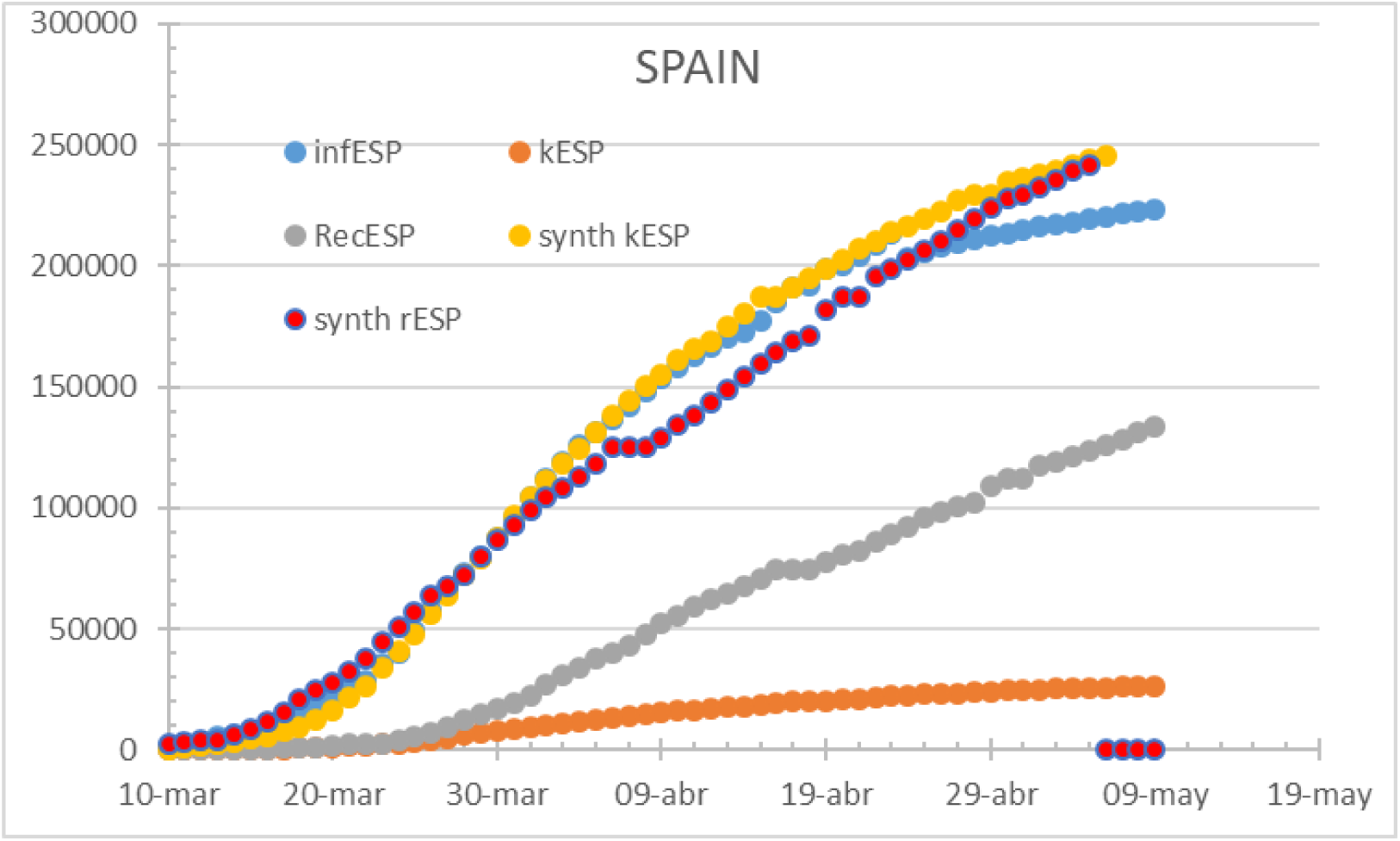
The updated accounting method triggers a gap between fatalities and unrealistically lowering infections.

The German case is also worth an analysis as it displays a consistent inconsistency since early April when the gap between detected infections and corresponding reconstructed fatalities mismatch. The country correlates an astounding HR of 97% with 10 days to fatality and 17 days to recovery curve.

**Figure 9.**
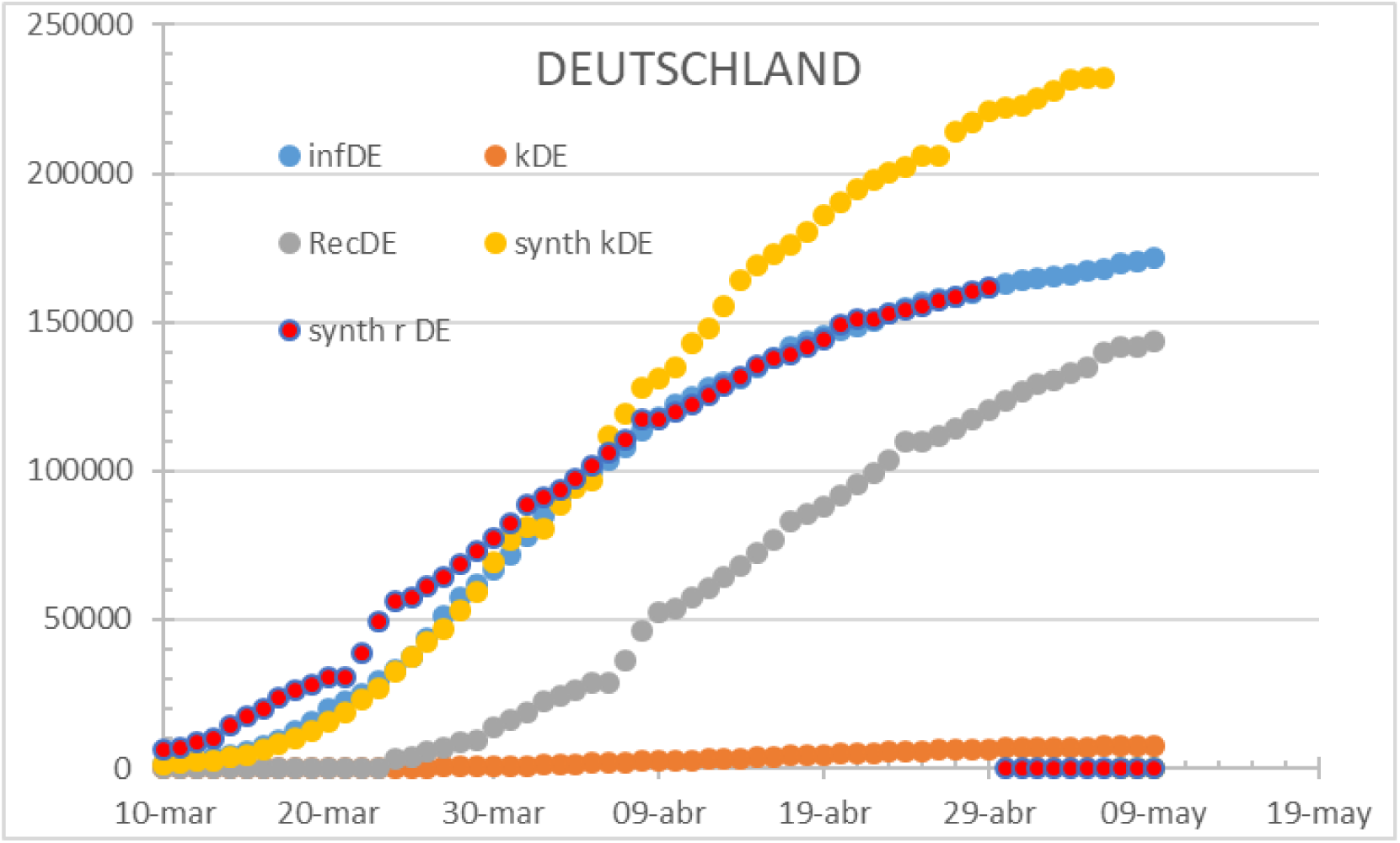
Germany should have around 250k infections and is reporting 70% of it as per may 14th. A more detailed analysis on the methodology changes since April 5th should be required to understand the mismatch.

South Korea has a Time to fatality of 7 days and a Time to recover of 25 days with a CFR of 1%. It displays data consistency until March 11^th^. After the date, official infections remain under the reconstructed curve from fatalities and above the recovered reconstructed curve. To remark that data form may 14^th^ is keeping the same inconsistency level. A note on the accounting and diagnostics could be of interest as the curves’ shape differs heavily and are potentially belonging to different causes.

**Figure 10.**
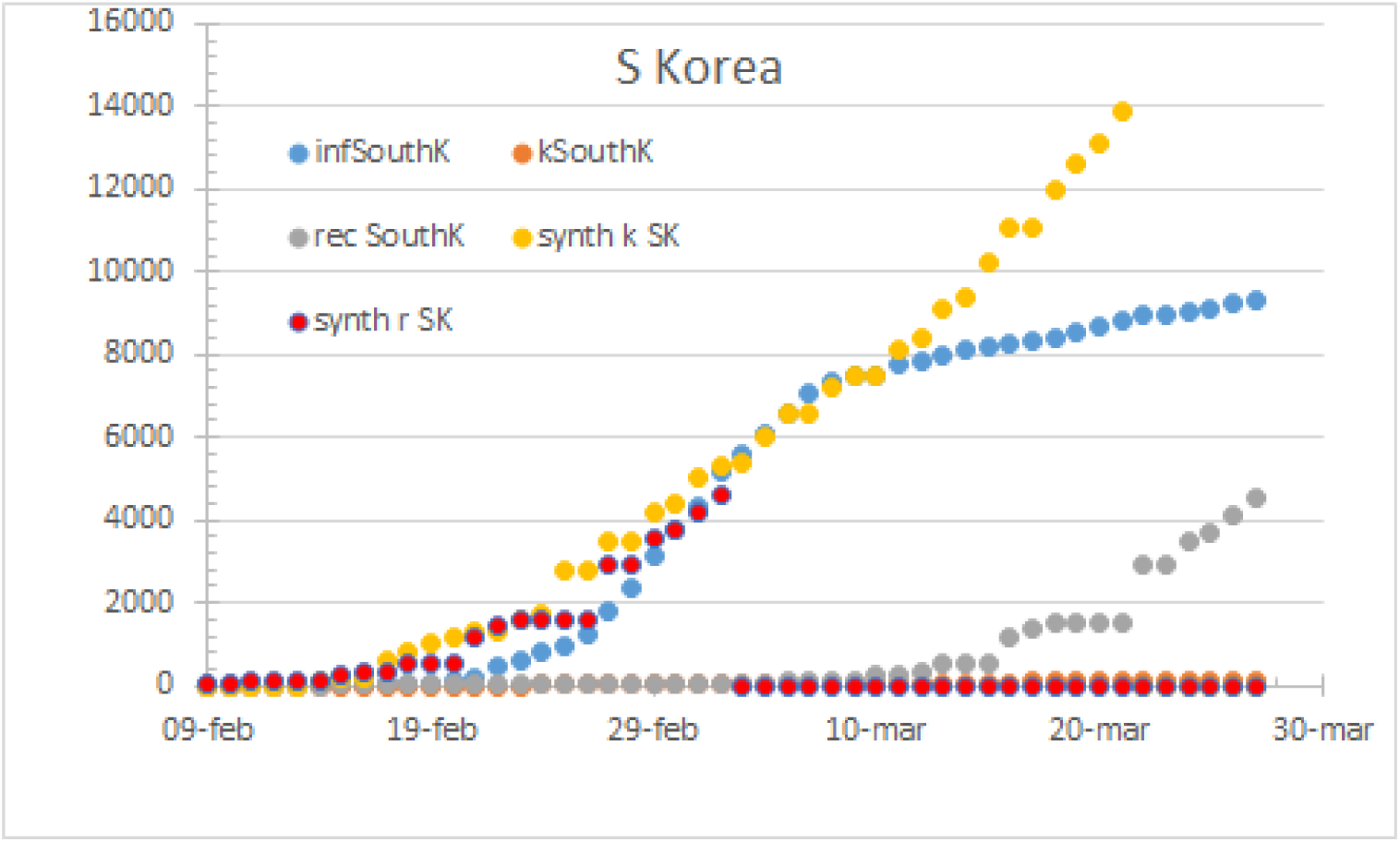
South Korea status by march 24th

**Figure 11.**
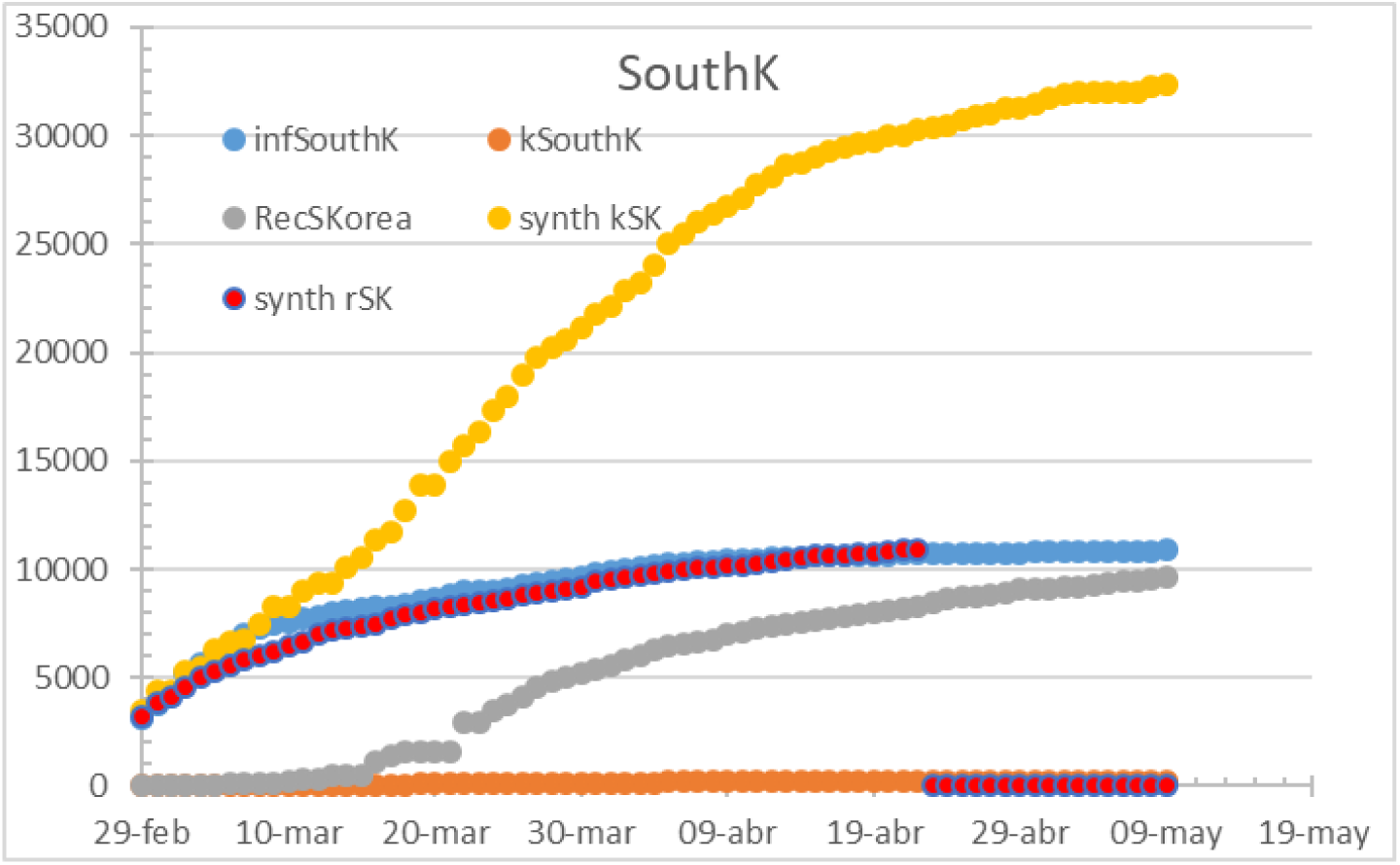
South Korea by May 14th. 91% HR, 4 days to fatality, 24 days to recovery.

Rest of the world has a Time to fatality of 6 days and a Time to recover of 16 days with a CFR of 7% on march 24th. The reconstructed infectious curves match the declared infections overall. By May 14^th^ the worldwide curves display a HR of 60% (matching infections), with 7 days to fatality, 20 days to recovery and a CFR of 8,9% however with an increasing gap between infections and fatalities, potentially due to better diagnosis and a limited control over COVID-19 spread.

**Figure 12:**
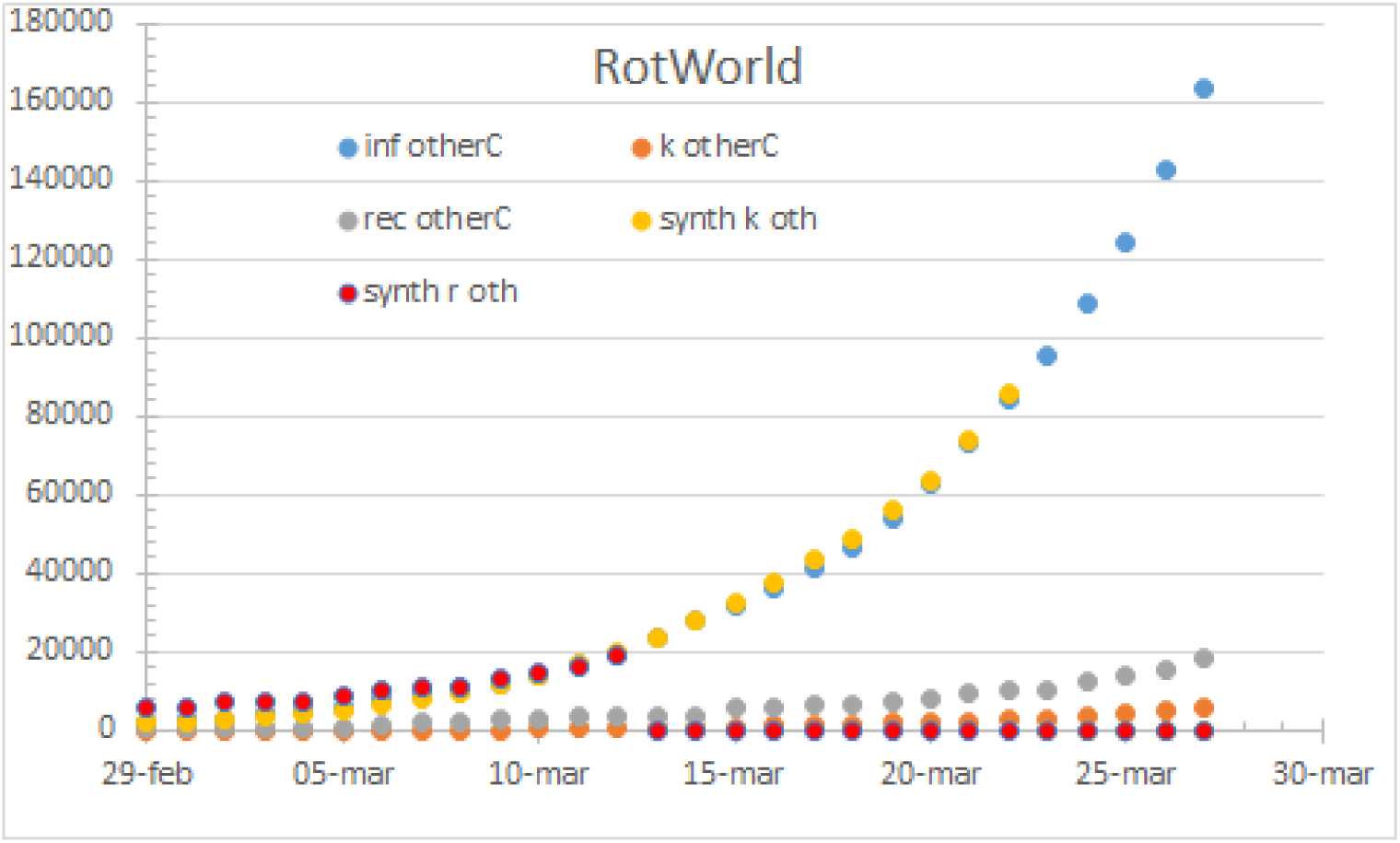
Rest of the world by march 24th. 100% HR

**Figure 13.**
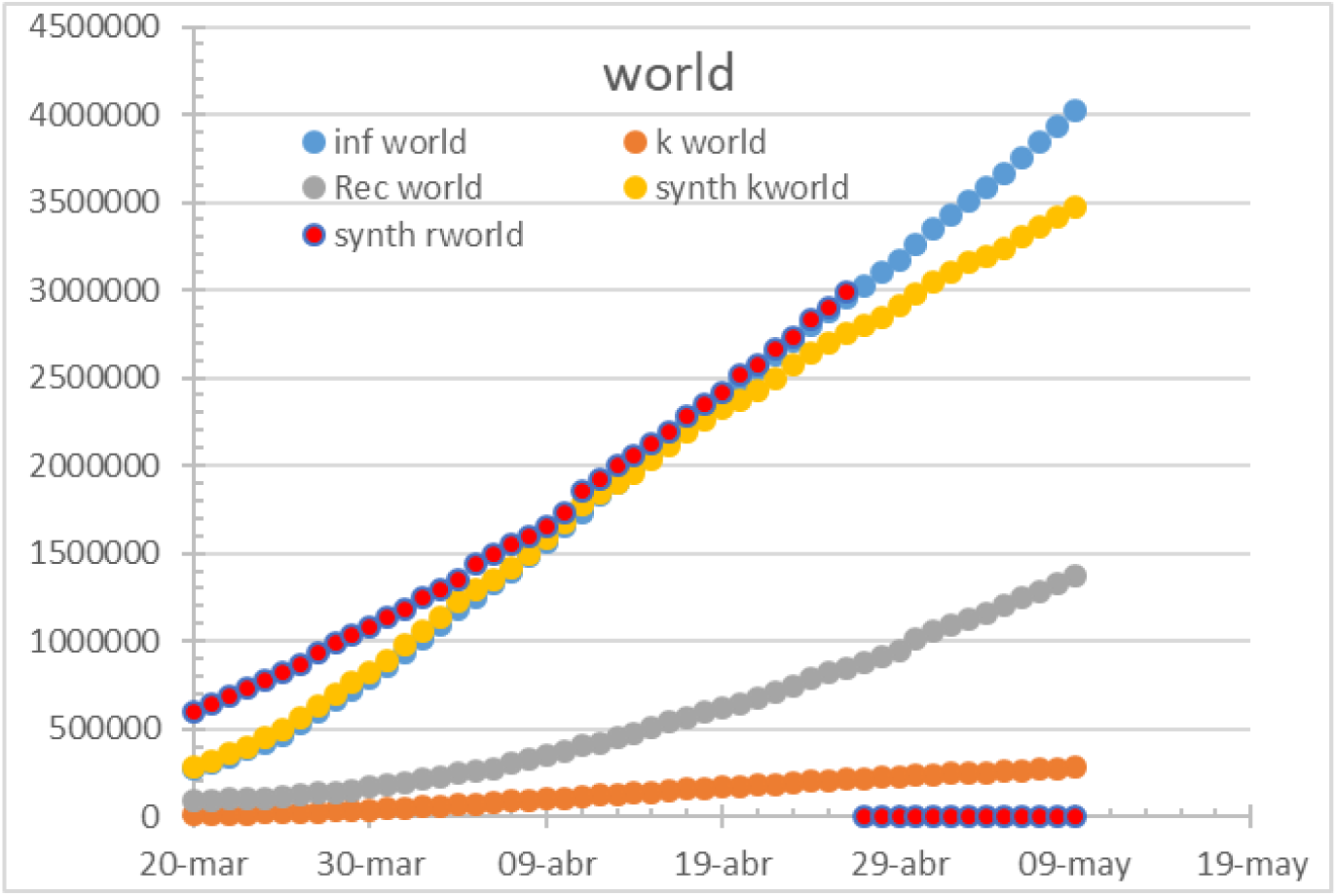
Worldwide status by May 14th.

The coefficients used to build up the reconstructed curves are respectively:

**Figure 14.**
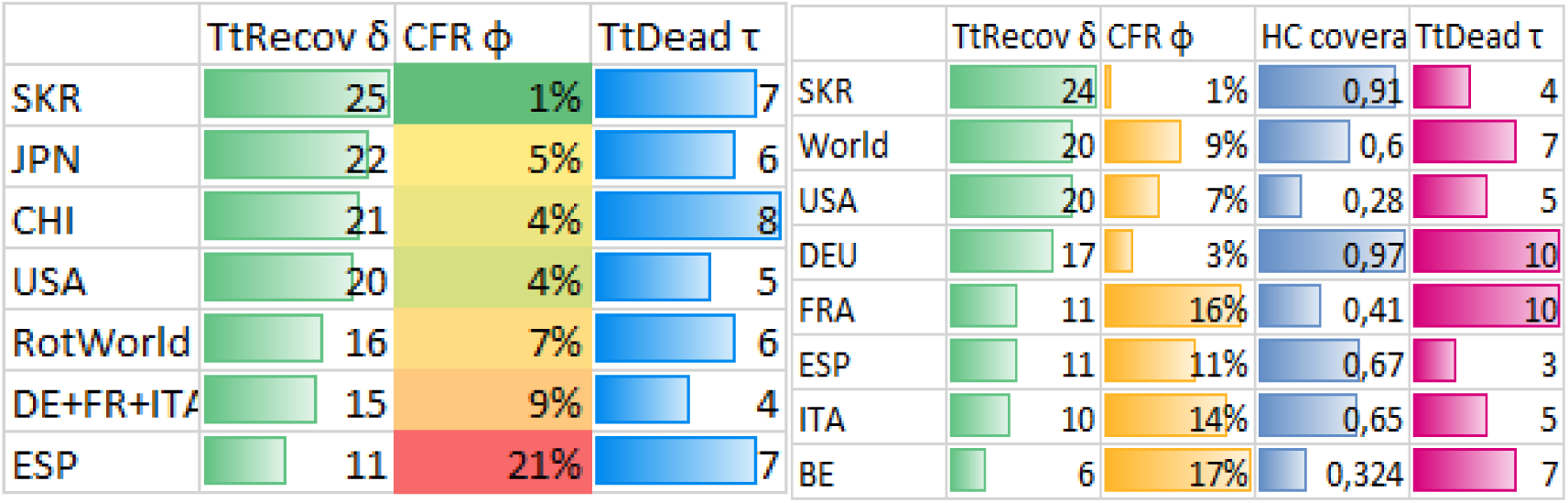
Coefficients used on March 24th -initial- and May 14th -updated- coefficients

The figures represent Time to Recovery as the time lapse from hospital admission of positive cases to hospital discharge and the Time to Fatality is the time lapse between hospital admission of positive cases to fatality record, and plotting the fatality rate against the variable Time to Recovery, the figure is generated:

**Figure 15.**
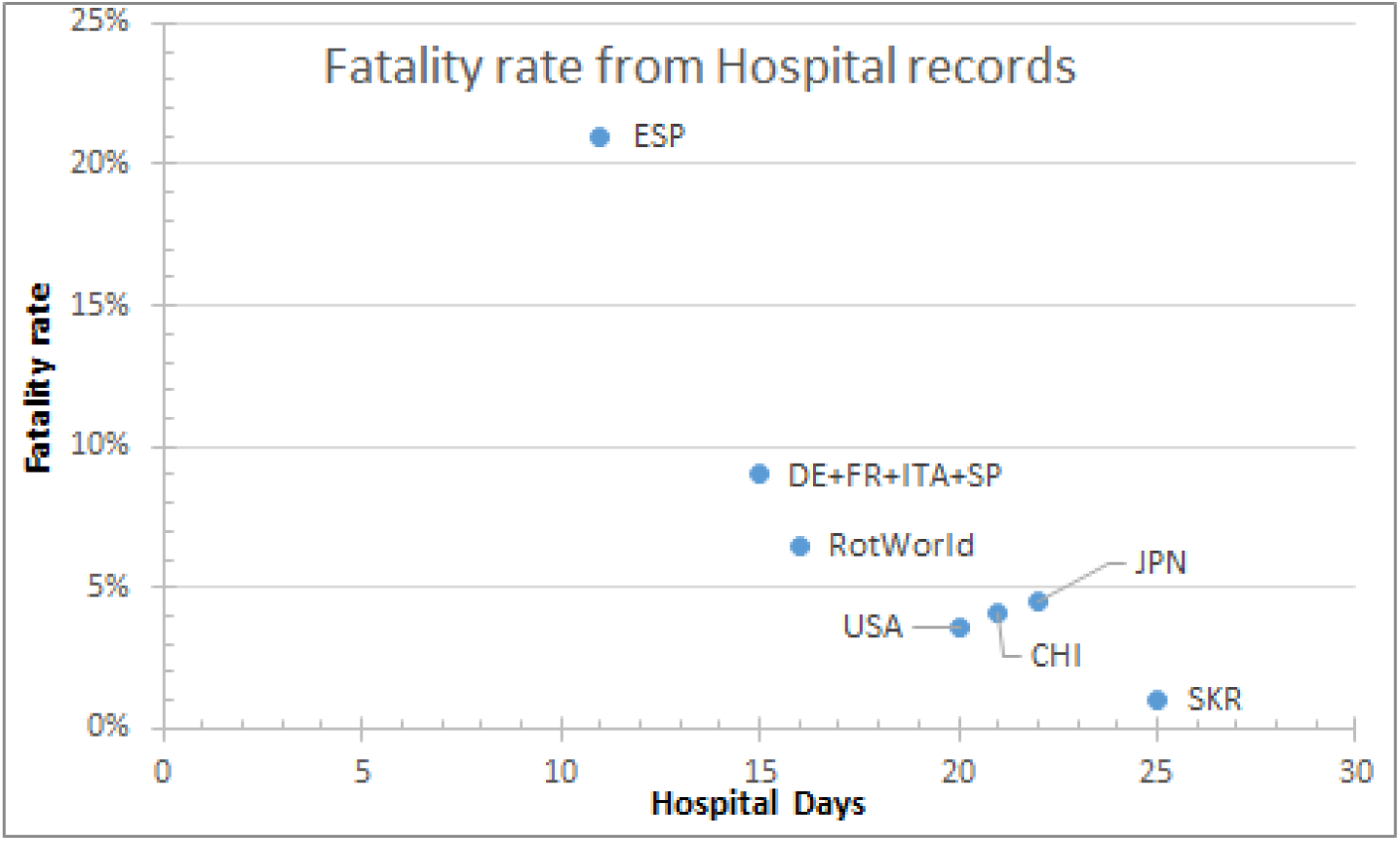
CFR Vs Recovery time by March 24th

**Figure 16.**
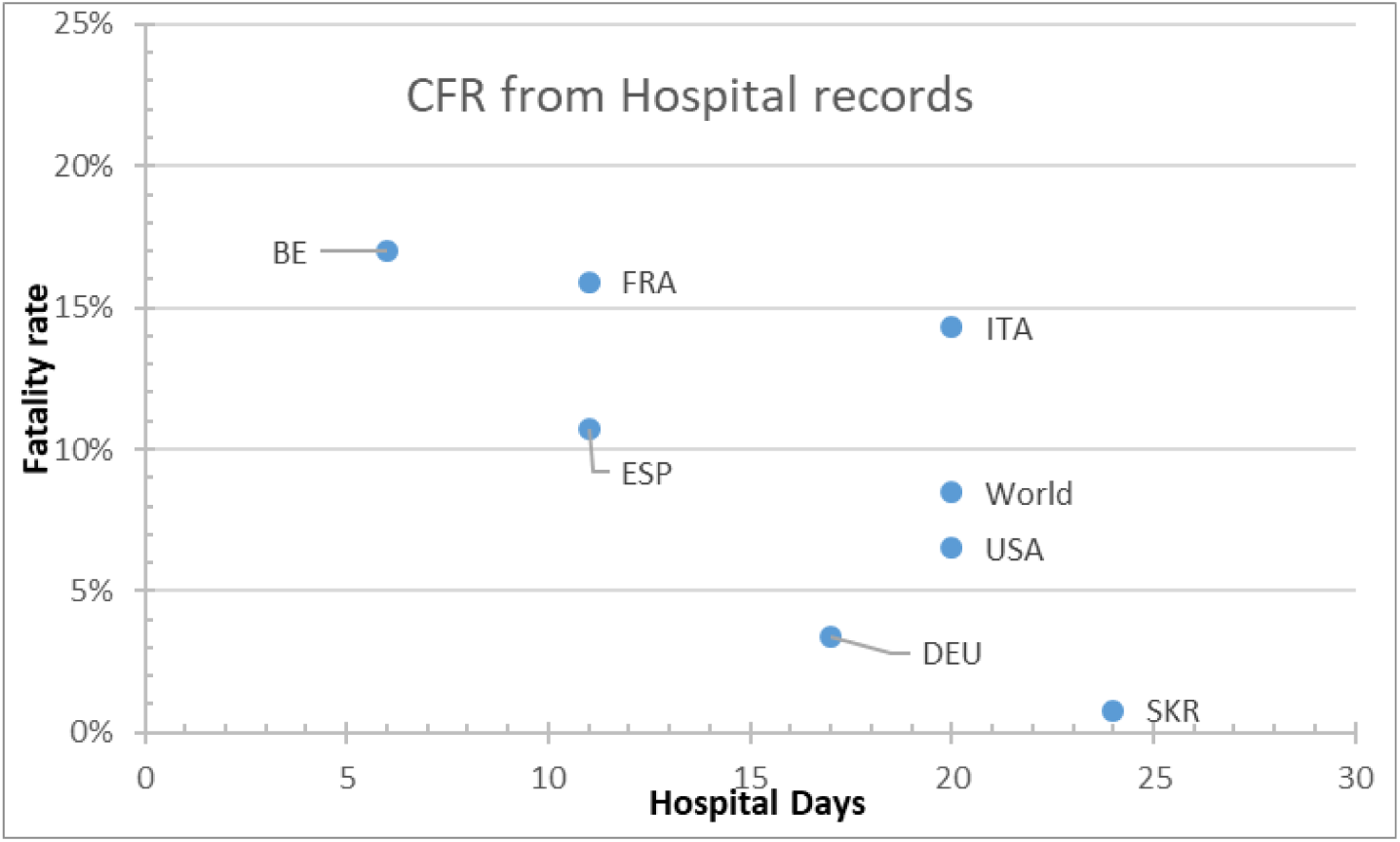
CFR Vs Recovery time bt May 14th.

Figures display that the fatality rate is lower as the hospitalization period is longer.

Applying the correction factor for the minimum estimation of infections on 2020-03-27 is presented on table.

| Countries | Minimum infections (estimation) |
| --- | --- |
| USA | 365.965 |
| CHI | 335.778 |
| DE+FR+ITA | 1.536.939 |
| ESP | 1.380.099 |
| SKR | 9.332 |
| Rest of the World | 1.065.448 |
| JPN | 6.606 |

## Discussion

Data provided from official sources is consistent both in Europe and the US as well as the rest of the world which reports on COVID-19 during its early stage. This indicates the suitability of data for the pandemic parametrization on initial data analysis.

In Asia, the Japanese procedures are accurate but induce delay in the signals and raw data must be corrected before modelling.

China has different curves from declared infection and the reconstructed from recovered and fatalities. The steeper curve belongs to the declared infection and hence, computing R0 on free flow from the declared infections curve may overestimate the real value of transmissibility if fatalities are well reported. Reconstructed curves are smoother and offer a consistent match which may get closer to real infection curves. The gap between declared and real infections can be represented as initial diagnostics were lagging the disease and once procedures were in place, the time to detection was increased, giving additional control over the situation.

The South Korean case displays a severe dissonance. While the fatalities curve shows a controlled-infection pattern (constant fatality rate) the declared infection points to a controlled stagnation pattern. Also, the underestimated recovery rate means that some COVID-19 positive patients are never discharged from the hospital, which is unlikely to happen.

The overall average time-to-fatality is of 7 days (time from anomalous infection symptoms leading to hospital acceptance, until fatality occurs in hospital premises), which remains in line with the published clinical course for COVID-19 and makes it consistent with ICU data.

Plotting hospital days (stay) Vs CFR shows that the mortality rate is much bigger in Spain, which is over an order of magnitude beyond countries as Japan, as China and the US which are in they turn well over South Korea’s CFR. In fact, what can be appreciated in the figure is that countries with completely saturated or unprepared healthcare system do experience a much higher mortality, potentially explained because overwhelmed Healthcare services are prone to decrease its patient stay and filter its admissions, focusing on the most critical ones. Patients are discharged faster and CFR is increased. Therefore, correlation between CFR and the time to recovery is not causal but explicative. Such an indicator can work for COVID-19 to measure efficiency on detections and national healthcare system overload. Roughly, the overall trend is to increase CFR by 1 point as the stay is shortened by 1 day. This estimator can be used to compute the effective number of hospital beds required to face a given pandemic infection or to determine the HC system capacity provided a fixed number of stations.

Therefore, the fatality rate against the hospital days curve is displaying the overwhelming of a healthcare system and it is not true to the real CFR of the COVID-19 unless we are near the X axis.

The following lines of the study will focus on the data and the parameter adjustment. The presented methodology for a first quality assessment demonstrates when data can be fed straightforward to a model in order to compute the epidemiological parameters and when the data requires preprocessing before feeding any realistic model or if the data is not even suitable for it, as the South Korea and German cases. Hence, anomalies as detected in South Korea, Germany and China potentially indicate that an evolved method to correct the baseline data must be applied to match consistently and understandably the curves with the reconstructions of such.

So far, CFR has to be considered a bad estimator for IFR (infection fatality rate) as the data is incomplete in many cases and the preclinical cases are unknown. However, the ratio from highest to lowest CFR can be a potential estimator on the real Infectivity where testing was not being conducted extensively Vs a full-population testing, providing a figure for total infected people at a given date, which can be contrasted with other methodologies.

## Data Availability

For the complete set of data (Including ancillary datasets) and elaborated files contact the corresponding author at. Latest data can be retrieved from the JHU at GitHub repository

https://github.com/CSSEGISandData/COVID-19/tree/master/csse_covid_19_data/csse_covid_19_time_series

https://cnecovid.isciii.es/covid19/#documentaci%C3%B3n-y-datos

## Contributors

OGR has developed and implemented the algorithms and made the data analysis and included corrections to feed epidemiological models. I must thank 4 anonymous reviewers for their valuable inputs on data interpretation.

And a special mention to the effort of the JHU for gathering and curating the datasets on GitHub.

## Declaration of interests

The author declares no competing interests.

## Acknowledgments

I have to thank all colleagues who helped me during the current study and the ones which proposed improvements and future research paths. I am also grateful to the many front-line medical staff who provided first-hand information and also for their outstanding and continuous dedication well before and beyond this outbreak.

## Notes

### Competing Interest Statement

The authors have declared no competing interest.

